# *One way or another, you’re not going to fit*: Trans and gender diverse people’s perspectives on sexual health services in the United Kingdom

**DOI:** 10.1101/2024.05.09.24307128

**Authors:** Tom Witney, Greta Rait, John Saunders, Lorna Hobbes, Laura Mitchell, Jay Stewart, Lorraine McDonagh

## Abstract

**Objectives:** Trans and/or gender diverse (T/GD) people in the UK are less likely to access sexual health services (SHS) than cisgender people but are more likely to report negative experiences. The British Association for Sexual Health and HIV (BASHH) developed expert recommendations for T/GD-inclusive SHS, but these lack service user perspectives. This study addressed this gap by asking T/GD people how SHS could be T/GD-inclusive.

**Methods:** Semi-structured interviews (n=31) and focus groups (n=21) were conducted with T/GD people aged 17-71 years old recruited through community organisations and social media, exploring experiences of SHS and inclusivity. Study design, materials, and analysis were informed by T/GD people and an advisory committee of charities and sexual health clinicians. Data were analysed using thematic analysis, managed using NVivo.

**Results:** Participants often expected that SHS were not set up for T/GD people. This was reinforced by poor experiences in other healthcare settings and the lack of information on NHS websites. Some participants had been denied care because they were ‘too complex.’ Participants wanted to know that SHS had engaged with the needs of T/GD people and looked for hallmarks of inclusivity, such as Trans Pride flags in reception areas. Some participants wanted specialist T/GD services, but others preferred to access general SHS. Staff attitudes were a key factor underpinning inclusivity. Anticipating having their identity questioned or needs dismissed, participants sought kindness and openness. Although the needs of T/GD people are diverse and different from cisgender service users, participants stressed that SHS staff already had the skills to deliver sensitive person-centred care and emphasised the value of inclusive SHS.

**Conclusion:** These findings provide insight into what a sample of T/GD people in the UK consider important for T/GD-inclusive SHS. Participants’ suggestions aligned with and reinforce BASHH expert recommendations. Importantly, they highlight the need for ongoing engagement to deliver T/GD-inclusive SHS.

**Key messages:** *What is already known on this topic:*

- Trans and/or gender diverse people are less likely to engage with sexual health services than cis-gender people
- The British Association for Sexual Health and HIV (BASHH) Gender and Sexual Minority Special Interest Group (GSM SIG) has developed expert recommendations for trans-inclusive sexual health services, but user perspectives are missing

*What this study adds:*

- Participant suggestions and preferences for inclusive services support BASHH GSM recommendations
- Participants looked for inclusive SHS that **recognise, understand and affirm** their needs

*How this study might affect research, practice or policy:*

- Enhancing T/GD inclusivity involves **active engagement** with **clinical spaces, processes and delivery**

## Introduction

Transgender is an umbrella term to describe people whose gender identity or expression differs from their sex assigned at birth. Gender diversity refers to the extent to which an individual’s gender identity, role, or expression differs from the cultural norms prescribed for a particular sex. This includes those who identify as transgender and those who do not identify within the traditional gender binary (1). Robust estimates of the number of trans and/or gender diverse (T/GD) people in the UK are lacking, however, the 2021 Census for England and Wales included optional questions about gender identity for the first time and 262,000 (0.5%) respondents indicated their gender was different to sex assigned at birth (2). The number of people recorded as identifying as transgender identity recorded in UK primary care records has increased fivefold from 2000 to 2018 (3).

T/GD people in the UK face high levels of violence, harassment, stigma, discrimination, marginalisation, homelessness and underemployment (4, 5). These are associated with increased rates of alcohol and substance use, anxiety, depression and suicidal ideation (6, 7). T/GD people face multiple, intersecting barriers to accessing healthcare across diverse settings (8-12). Lack of information about sexual and reproductive health (SRH) needs of T/GD people, among both service users and providers, is a barrier to good quality care (13). Multiple structural, community, network and biological factors are associated with greater risk of HIV acquisition and lower rates of HIV testing among T/GD people globally (14-16). A survey of T/GD people in the UK suggested they are less likely to use sexual health services (SHS) than cisgender people (16). T/GD people using online sexual health testing had higher rates of HIV and STI positivity compared to cisgender service users, and reported complex sexual health needs, including engaging in chemsex, group sex, fisting and sex work (17). However, data are inconsistent and the first estimate of HIV prevalence among T/GD people in England suggested a similar prevalence to the general population (18).

The British Association for Sexual Health and HIV (BASHH) Gender and Sexual Minority Special Interest Group to produced expert recommendations for how sexual health services can provide inclusive care for T/GD people (19). The authors highlighted the lack of published evidence, including a need to understand service user perspectives. The aim of this study was to explore T/GD people’s experiences in SHS, and perspectives on how best to support their sexual health needs.

## Methods

### Design

This qualitative research was conducted by a cis-gender research team, guided by input from an expert steering group, consisting of representatives from trans community organisations and sexual health clinicians, and from T/GD patient and public involvement (PPI) representatives. The project was informed by published recommendations for research with trans participants (1). Participants were offered a choice of an individual interview or participating in a focus group. Ethical approval was granted by University College London Research Ethics Committee (8805/007).

### Participants and recruitment

Participants aged ≥16 years old who self-identified as transgender and/or nonbinary and living in the UK were eligible to participate. Potential participants were invited to register their interest in the study via T/GD community organisations, social media, word of mouth and snowball sampling.

Purposive sampling was used to ensure representation of different gender identities across the sample. Participants gave informed consent to participate in the study.

### Procedure

Interviews lasted 45-90 minutes and were conducted via Zoom by an experienced researcher (TW, he/him), guided by a topic guide (on-line supplementary document 1). Interviews explored participants’ experiences of accessing SHS and their reflections on the inclusivity of services. Focus groups lasted 90 minutes and were conducted via Zoom by TW, supported by a research assistant. Discussions focused on participants’ shared understanding of sexual health, three patient vignettes and reflections on an ‘ideal’ T/GD-inclusive SHS. Both interviews and focus group discussions were digitally recorded and transcribed with informed consent. A voucher of £50 was offered to each participant. Data collection continued until the team judged that sufficient information power had been generated to meet the research aim (20).

### Analysis

Data were analysed by TW using thematic analysis (21), supported by NVivo. Data were coded inductively and developed into initial themes. These themes were refined through iterative engagement with transcripts and named using data extracts from interviews and focus groups. Themes were validated by discussion between the research team (LMD, she/her; JSa, he/him; GR, she/her) and in consultation with steering group members (LH, she/her; LM, she/her; JSt, he/him) and PPI representatives.

## Results

Two-hundred and ninety-six people expressed an interest in participating in the research, of whom fifty-nine were recruited to the study (see Table 1). Thirty-one individual interviews and three focus groups (n= 8; n=8; n=6) were conducted on-line between May and July 2022. A low number of trans women/transfeminine people initially expressed an interest in participating. To address this shortcoming, snowball sampling was used to recruit an additional focus group with trans women (n=4), which was conducted in July 2023. Participants reported diverse gender identities, sexualities and relationship types (see Table 2).

**Table 1:**
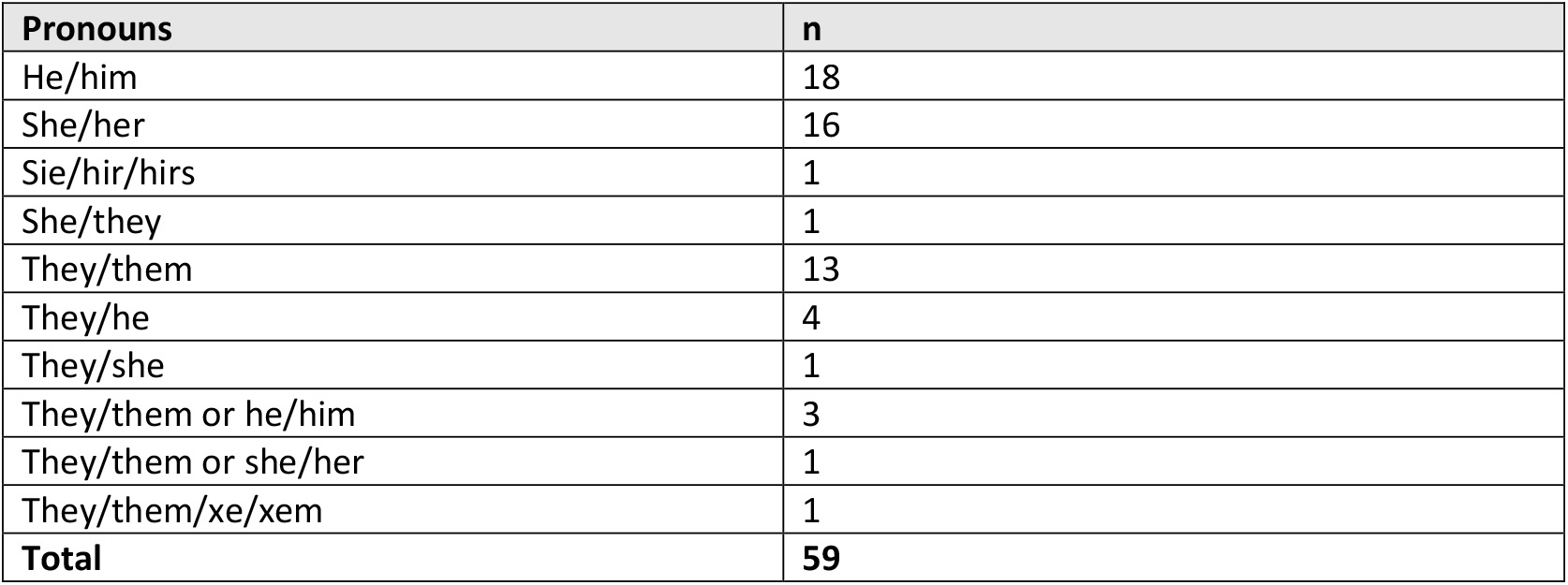
Participant summary - pronouns.

**Table 2:** Sociodemographic details (provided by 53 participants)

| Age |  |
| --- | --- |
| 17-24 years old | 17 |
| 25-34 years old | 24 |
| 35-44 years old | 5 |
| 45-54 years old | 5 |
| 55-64 years old | 0 |
| 65 years or older | 1 |
| No answer | 1 |
| Ethnicity |  |
| Black/black British | 2 |
| Indian/Indian British | 3 |
| Mixed | 5 |
| White/white British/white Irish/white Welsh | 43 |
| **Sexuality** |  |
| Bisexual | 14 |
| Demisexual | 3 |
| Gay man | 7 |
| Gay woman/lesbian | 3 |
| Heterosexual | 6 |
| Pansexual | 8 |
| Queer | 12 |
| **Current relationship\*** |  |
| Dating | 13 |
| Engaged | 3 |
| Married/Civil Partnership | 6 |
| Monogamous | 9 |
| Open | 7 |
| Polyamorous | 9 |
| Separated/divorced | 2 |
| Single | 17 |
| **Education** |  |
| No qualifications | 1 |
| GCSE, or equivalent | 4 |
| A-levels, or equivalent | 11 |
| Foundation degree, or equivalent | 4 |
| Honours degree, or equivalent | 14 |
| Masters degree, or equivalent | 15 |
| Doctorate, or equivalent | 4 |
| **Employment\*** |  |
| Employed full-time | 24 |
| Employed part-time | 8 |
| Employed flexibly | 4 |
| Full-time student | 12 |
| Part-time student | 3 |
| Not in education or employment | 9 |
| Retired | 1 |
| Self-employed | 4 |
| **Location** |  |
| Urban (population >10,000 people) | 45 |
| Rural (population <10,000 people) | 8 |
| **Disability** |  |
| Yes | 26 |
| No | 27 |

\*Respondents could select more than one option

Data were coded and organised into initial themes: barriers to inclusive care, facilitators of inclusivity and how participants characterised inclusivity. From these, three cross-cutting narrative themes and subthemes were developed, which are presented here. Full supporting quotations are provided in Table 3.

**Table 3:** Themes and illustrative quotes.

| Theme/subtheme | Description | Quotes |
| --- | --- | --- |
| ***“One way or another you’re not going to fit”*** |  |  |
| An expected lack of inclusivity | Participants expected services not to be inclusive or to be discriminatory | Services have inherently cis-normative, hetero-normative assumptions [...] obviously as a trans person you are most likely in one way or another - by virtue of being trans - not going to fit<br>Focus group participant (they/he) |
|  |  | It’s scary by definition. The fear is that you are going to be treated badly. You’re going to be treated as something different rather than just an individual with those concerns, those problems, you know<br>Interview 14 (she/her) |
| Poor experiences in other services | Experiences in other NHS services lowered participants’ expectations of care from sexual health services | I think the thing about trans healthcare in this country that is necessary to understand as a first principle, is that everything is on fire [...] The assumption facing a trans person from the moment they enter the system as a patient, is that they are somehow a problem<br>Interview 17 (he/him) |
|  |  | [Waiting a long time for gender services] does colour how you see the rest of the NHS, because you’re getting the message that nobody cares about me, it’s fine for me to wait five years for care, for something that’s really important to me. So you do tend to think that the rest of the NHS will treat you like shit as well, and have a long waiting list, misgender you all the time. It doesn’t improve patient relations with sexual health services, or anybody else<br>Interview 20 (he/him) |
|  |  | It's not a secret that no one in this country knows fucking anything about trans healthcare because it's literally the biggest fucking mess. So when it comes to trans stuff, a healthcare provider is a means to an end. I'm not always honest with them; I will do what I have to do to get the help I need from them and I don't expect them to be, necessarily, on my side in seeking that help"<br>Interview 25 (they/he) |
| Denial of care | Some participants reported that trans and gender diverse people are routinely denied sexual health care | If we disclose our trans status when seeking sexual healthcare, we will immediately be denied, like before anything else. It's either, "You're too complicated to deal with, you need a specialist," or "Hmm, yes, you do have raging thrush and do you think that that's probably because of your hormones and you should de-transition about that."<br>Interview 17 (he/him) |
| "Uphill struggle" | Participants described issues with being able to access interventions that are routinely offered to other demographics | I had a great deal of trouble accessing PrEP because they don't offer PrEP to heterosexual cisgender women and it took me quite a while to convince them, on the phone, that that was not me... the service is not accessible in that way unless you fit their expectations.<br>Interview 24 (she/her) |
| The NHS doesn't understand T/GD people | The lack of appropriate information led some participants to question if the NHS understood T/GD people | It's very 'these are the body parts a man has, and these are body parts a woman has.' And if you're trans you're going to have to ask for something different<br>Interview 4 (he/him) |
|  |  | Any time I want to look up something [on the NHS website] related to body parts I have it's always "women sometimes blah, blah", and I have to actively push past that to absorb the information when it's misgendering me as I'm reading. [...]<br>There are days when I genuinely don't care but other days it does impede me processing the information when I have to read the word woman in every paragraph<br>Interview 28 (he/him/they/them) |

| ***"If people come in, they shouldn't be a surprise"*** |  |  |
| --- | --- | --- |
| Specific T/GD inclusivity | In the context of increasing hostility towards T/GD people, participants sought indications that services would be specifically inclusive, such as a trans pride flag displayed in the service | It's nice to see [the trans pride flag] but you see it and you think, OK, this organisation probably isn't going to be transphobic. And, I think just people being explicit about their trans inclusivity because, sadly, you know, simply having a generic Pride flag or saying we're LGBT supportive, that's not really the comfort anymore that it necessarily should be.<br>Interview 12 (she/her) |
|  |  | The flag always helps but not just the generic rainbow flag because we know the NHS have taken that and that makes it ambiguous but the Progress flag, or the trans flag. Or something like it being visibly there on the website, or saying we cater to LGBT people or something like that, just making it very specifically clear that it is friendly.<br>Interview 29 (he/him) |
|  |  | Having a trans flag, because you can see, at least they're doing a little bit. So they've thought about it as opposed to not having thought about it at all.<br>Interview 04 (he/him) |
| Gender neutral greetings | Being greeted by staff in a gender-neutral manner made a significant different to participants | That's the difference between me feeling comfortable and uncomfortable. I don't think the average man or woman is bothered whether they get called sir or madam at a clinic. Just say, hello, that's it.<br>Interview 3 (she/her) |
| Physical spaces | The arrangement of physical space, including waiting rooms, and provision of gender-neutral toilets helped participants feel welcomed in services | Oh my god, they had like women over there, and men over there... and I don't know what I would do now if I was going to that clinic like being trans<br>Interview 20 (he/him) |
|  |  | With toilets, you know, inaccessibility to these toilets, kind of, going into a service and it has a men's and a women's, and it's like already you're fucking things up a little bit here.<br>Interview 11 (she/her) |
| Registration forms | Having inclusive options on forms helped build confidence that services were engaged with the needs of T/GD people | Looking at a form and is it non-binary inclusive, because even though I'm not non-binary if I know that they're being non-binary inclusive I know that they've thought about trans men as well<br>Interview 04 (he/him) |
| Sharing pronouns | Staff sharing their pronouns, or wearing pronoun badges gave T/GD service users confidence care would be inclusive | I find [it] easier when the other person introduces their pronouns. That makes that "OK now I can" because I often am scared that these health providers will have their own biases<br>Focus group participant (they/them) |
| Service configuration | Participants expressed divergent opinions on whether services should be T/GD-specific or whether they would prefer inclusive general services and acknowledged that one size would not fit all | Having specific clinic times set aside for trans-people so that you're not waiting in a room and worried about other people looking at you and going "who's that?"<br>Interview 21 (they/them/he/him) |
|  |  | Essentially, I'm with like-minded people [...] OK no one in this waiting room is looking at me as if I'm a freak of nature walking through the door, which just makes the whole process just feel a little bit nicer<br>Interview 11 (she/her) |
|  |  | In an ideal world, everything is just sexual health care and it's normalised for everyone. If you have more specialised [T/GD] clinics, maybe you have some lovely people lurking outside that want to harass anyone that goes in<br>Focus group participant (she/her) |
|  |  | You know, how insane the wait list for the GIC is, [T/GD specific clinics] could just end up being a repeat of that<br>Focus group participant (she/her) |
|  |  | It would just be nice if things that are good for trans people didn't have to be so kind of portioned off from everything else [...] you know, 10 years ago I wouldn't go into a waiting room if I knew that it meant that being sat in there meant that everyone in there knew I was trans... I think when people try and be accessible they need to consider that that in itself is |
|  |  | actually a hurdle for some people and that it's good to kind of mix things up a bit so, that there's options for everyone<br>Interview 27 (he/him) |
| ***"You can talk to people normally"*** |  |  |
| Expectation of good care | Participants expressed a desire for all services – even non-specialised ones, to provide good quality, T/GD-inclusive care | Just having confidence that we're going to get the same sort of treatment and good treatment from whoever that it is that we approach because everyone is confused, and that's not good.<br>Focus group participant (he/him) |
|  |  | It would just be nice if things that are good for trans people didn't have to be so kind of portioned off from everything else<br>Interview 27 (he/him) |
| Staff openness | Part of inclusive care for participants was staff having an openness towards T/GD service users' needs and to learn from them | It's very much about just doctors in general how they present themselves to you, you're more likely to respond if they seem open and not like they're going to judge you<br>Interview 6 (they/he) |
|  |  | Just that you have that connection with [staff] – you can just relax in the situation and you don't feel they're going to find you odd, or not really understand you, or you're just not on the same page [...] And an openness, like a curiosity but not like an over curiosity but like... kindness and understanding<br>Focus group participant (they/he) |
| Educating staff | Participants were often ambivalent about educating staff about T/GD people's sexual health needs | It's unfortunate that it's trans people attending these services that have to teach the service providers how to do their job properly, but sometimes that is how it goes.<br>Interview 18 (he/him) |
|  |  | I am patient with people because they're not asking out of malice, they generally genuinely want to know... The harm has already been done to me, if I educate these people now it means that the next person who comes in and is in my position isn't made as uncomfortable<br>Focus group participant (they/them) |
| Specialisation | While participants did not expect all staff to have a detailed knowledge of T/GD people's sexual health needs, they wanted access to expertise | I don't expect every sexual health professional to be fully up to date with all the latest trans-specific stuff, I know that we're a niche group. But for there to be some very explicit ones to go to, maybe if a certain clinic has a worker on staff who maybe specialises in this sort of thing<br>Interview 03 (she/her) |
|  |  | I'm sure there are some really good professionals out there when it comes to sexual health and gender diverse people. But I think you would still rather start with someone you know gets it<br>Interview 03 (she/her) |
| Inappropriate curiosity | Participants experienced staff asking inappropriate questions about their identity that were not related to the provision of care | Last time I was in for a sexual health check-up, the person who was asking me all these questions about what surgeries I had had, which is fair enough I can excuse that because that might be relevant to later on in the consultation or whatever, but then was asking me all these questions about what surgeries I have got planned to have, and what's down the road for me, and why is it hard for trans people to come to sexual health clinics, and why does that cause dysphoria and why does that make you uncomfortable and how did you know that you were trans, and when did you find out, and how old were you? And that's pushing the professional role too far.<br>Focus group participant (they/them) |
| Inclusivity means being treated normally | For some participants, being treated carefully by staff was a source of frustration | I guess it's trying to remember that you can kind of just speak to people normally [...] I get that it comes from a place from not wanting to upset me but then I almost find it frustrating because I'm like... this conversation is just taking longer!<br>Interview 27 (he/him) |
| Cis/heteronormative assumptions | Participants described how assumptions were made about their genitals and sexual practices when receiving sexual healthcare | If I put in that I was a man and that I had sex with men I would get a throat swab, a bum swab, and a pot to piss in – there was no way for me to communicate that I needed the [vaginal] swab not the pot... eventually they'd figure it out... and they would just hand me back a single swab... |
|  |  | Interview 27 (he/him) |
|  |  | When I started on PrEP, they were like, “You can do the event-based PrEP”, and then decided that I couldn’t [...] based on nothing other than their own preconceived notions of how they think that a trans person of my type should have sex<br>Focus group participant (they/them) |
|  |  | Things can be a bit one size fits all unless you ask for it to be different [...] when actually what they could do is be asking about body parts. So, these are questions we ask everybody which of these types of sex do you have [...] So there’s ways of asking it, do you have sex with people with penis, people with vagina, you know, doing it that way, more like an inventory. And then from there that could inform what labels get printed out for you or what testing kit gets sent to you, rather than just these very, sort of, non-binary exclusionary quite frequently”<br>Interview 04 (he/him) |
| Person-specific terminology | Having the opportunity to share the terms they preferred to use for their bodies was important for some participants | If you’re doing like the list the contents of your abdomen thing you can also just put next to it like preferred language for this thing, so just have a little write in... Please don’t refer to this as a vagina, please just say this<br>Focus group participant (he/him) |
|  |  | I don’t like to use female terminology for my body so I would say I have frontal sex or the front hole or instead of the term vagina, I really hate that word it makes me feel uncomfortable<br>Interview 29 (he/him) |

### 1. “One way or another, you’re not going to fit”

Participants often believed that SHS were not set up for T/GD people. In some cases, this was based on negative experiences in other healthcare settings. Some of those who did engage with SHS described being denied care, either on the grounds of complexity, or because of ‘trans broken arm syndrome’ (22) where being transgender is inappropriately suspected as causing a complaint. Other participants portrayed an ‘uphill struggle’ accessing risk reduction interventions, such as HIV pre-exposure prophylaxis (PrEP).

Those who sought information about sexual health on the NHS website described how the lack of T/GD specific content meant the information was “*aimed at the exact opposite people that I am*” (Interview 1, she/her). Not only was this dehumanising and upsetting, it made some participants question whether the NHS acknowledge the existence of T/GD people. Participants also wondered if SHS essentialised T/GD service users as the sex they were assigned at birth, for example that trans women are “*people who have sex with men but who are, in the eyes of doctors, also kind of a man*” (Focus group participant, she/her).

These experiences, both of SHS and wider healthcare, compounded by experiences of societal transphobia, contributed to low levels of engagement with SHS among participants and shaped the expectations of those who did engage with services.

### 2. “If people come in, they shouldn’t be a surprise”

To overcome expectations of a lack of inclusivity, participants wanted to know that SHS had engaged with the needs of T/GD people. In the context of increasing hostility towards T/GD people, some sought reassurance that they would be welcome, “*you don’t always have that assurance that [LGBT spaces] are going to be trans friendly*” (Interview 11, she/her). Having Trans or Progress Pride flags in reception areas indicated active engagement with T/GD inclusivity. Some participants felt a statement of inclusivity was a sufficiently positive indicator but for others having explicit T/GD policies supported their trustworthiness, “*describing on their website exactly the things that they do to make them trans-inclusive; not just saying that they’re trans-inclusive*” (Interview 21, they/them/he/him).

Some had experienced being misgendered when arriving at SHS. In services where reception staff avoided using gendered terms when greeting people, this simple change had a positive impact. The arrangement of physical spaces and facilities was also important for a sense of being expected and welcomed by services, for example not having waiting areas segregated by gender and provision of neutral toilets. Registration forms with appropriate gender options were also an indication of inclusivity, and having non-binary options further increased confidence that services were engaged with the needs of T/GD people. As well as indicating inclusivity, registration forms could also help facilitate consultations (see ‘*You can talk to people normally’*, below). Staff sharing their pronouns at the start of consultations or on name badges could help establish that they were engaged with the needs of T/GD people.

Participants had diverse opinions on how services should be configured. Some would prefer a dedicated clinic, feeling more comfortable in waiting areas with other T/GD people, but expressed concerns that this could make it a target for harassment or funding cuts. Others preferred being seen as part of general services, to support a sense of normality and to avoid ‘outing’ themselves.

### 3. “You can just speak to people normally”

Interactions with staff during consultations was a key factor that underpinned good experiences of care. Anticipating having their identity questioned or needs dismissed, participants sought kindness and openness. Participants recognised that sometimes staff with less experience of providing care to T/GD people were anxious not to cause offence. While the majority appreciated the effort being made, some found it frustrating, “*I get that it comes from a place from not wanting to upset me but… this conversation is just taking longer!*” (Interview 27, he/him).

Participants emphasised the importance of avoiding making assumptions about T/GD people’s genital configurations or sexual practices during interactions, for example when providing kits for self-sampling or discussing event-based PrEP. They suggested that registration forms that gave service users the opportunity to share an ‘organ inventory’ could help facilitate this. Some participants had terminology they preferred to use for their bodies, for example ‘front hole’ instead of ‘vagina,’ and having the opportunity to share these preferences at registration and have them used during consultations was an important way for them to be affirmed in the interaction.

Participants did not expect healthcare professionals to have a detailed knowledge of T/GD people, but to have a ‘*general awareness’* (Focus group participant, they/he) of their sexual health needs. Although some felt it was important for them to teach clinicians about T/GD sexual health during consultations, others described situations where they had been made uncomfortable by intense questioning about their identity or about topics not relevant to the conversation, for example about plans for affirming surgeries. Participants suggested that staff should be aware of and proactively offer additional services that would support T/GD people, including provision of cervical smears for those with a cervix; monitoring for those self-sourcing gender-affirming hormones and T/GD appropriate support for people who had experienced sexual assault.

Although the specific needs of T/GD people might be diverse and different from cisgender service users, participants stressed that SHS staff already had the skills to deliver sensitive person-centred care: *“you don’t have to build from the ground up in terms of knowing how to get people to talk about their bodies and their health problems*” (Interview 17, he/him). Several participants who had positive experiences wanted to emphasise the value of inclusive SHS: “*I feel very safe there. I feel listened to. And I don’t feel awkward or an anomaly or strange, or other. That’s really, really special in healthcare”* (Interview 029, he/him).

## Discussion

Compounded by experiences of societal transphobia, participants’ negative experiences of healthcare contributed to low levels of engagement with SHS and shaped the expectations of those who did engage with services. This study found that T/GD people often expect that SHS are not designed to meet their needs, that they look for signs when judging if a service is inclusive and that a person-centred approach to consultations, along with basic awareness of T/GD people’s sexual health needs, can help to meet common expectations of good care. These findings highlight how simple changes can impact on T/GD people’s experience of services. The lack of consensus among participants suggests that one model will not be sufficient to meet the needs and preferences of all and the need for inclusive practice in all services.

The strengths of this research are its inclusion of participants with diverse gender identities and the involvement of a PPI group throughout the research to ensure questions, approach, interpretation were appropriate. Through the development of narrative overarching themes, this analysis is specific to sexual health and goes beyond barriers and facilitators of access to healthcare, which have been well explored in other studies (8-12). The limitations of this research centre on the predominantly White, urban, university educated sample (see Table 2) which means that the experiences and perspectives may under emphasise the role of intersecting inequalities and limits transferability to other settings.

The findings offer support for the recommendations of the BASHH Gender and Sexual Minority Specialist Interest Group regarding trans-inclusive sexual health services. These recommendations emphasise the need for gender-neutral registration forms, waiting and examination rooms, as well as toilets. Additionally, they underscore the importance of staff undergoing proper equality and diversity training. From a clinical perspective, the recommendations focus on several key aspects, including asking and respecting patients’ pronouns, tailoring STI testing to individual sexual practices and risks. Furthermore, the recommendations encourage inquiring about experiences of interpersonal domestic and sexual violence, recognising that these are common challenges faced by T/GD people.

Despite the existence of the BASHH recommendations, the widespread low expectations of SHS among participants emphasises the need for services to overcome these perceptions and engage with T/GD people. The importance of community in engagement with healthcare has been stressed in other studies with T/GD people (23). This points to the importance of developing trust and establishing the trustworthiness of services as part of this process. Research on physician patient relationships point to the role that social and historical context can play in mistrust between people from marginalized groups and healthcare services, as well as the importance of respect, partnership and time and consistency in building trust (24, 25). Other studies of LGBTQ+ experience of services describe how physical spaces, service infrastructure and interactions with staff are all critical to creating “safe spaces that matter” (26). Allyship for healthcare professionals includes embracing new ways of thinking and delivering care to those who might typically be excluded (27). T/GD people in this research had diverse sexual health needs, linked to not only their identity and gender affirming care, but also their relationships and sexual practices, which changed over time for many. However, by following a person-centred approach to consultations, the needs and priorities of each individual can be ascertained and met. National guidelines on person-centred sexual history taking were updated to reflect gender diversity in 2019 (28). The diversity of sexual health needs are linked to gender affirming care, which is a critical component of the sexual and reproductive health of T/GD people (29).

These findings help to provide an evidence base for the BASHH expert recommendations and provide clinicians with some very simple and easy to adopt recommendations that make a significant impact to T/GD service users, such as ‘just saying hello’ and including pronouns when introducing themselves. They also outline structural changes that could be made to facilitate easier interactions, for example on registration forms, electronic patient records, and self-sampling kits. However, they also highlight the challenges of providing services that will meet everyone’s preferences but provide principles of inclusivity that can be delivered in all services.

Future research on provider perspectives in UK SHS would complement emerging research in other healthcare settings (30). These findings could be used to inform the development and implementation of interventions to improve the experiences of T/GD people in UK SHS. They could also inform the development of further research to quantify how common these experiences are and develop understanding of how they are associated with sociodemographic and behavioural variables.

## Conclusion

These findings are consistent with and provide further support for existing BASHH GSM recommendations for inclusive SHS. They highlight the need for services to actively engage to reach a population with potentially greater sexual health needs who are disengaged with services. To meet needs of range of T/GD people there is a need for both specialised clinics and general inclusivity of services.

**Figure 1:**
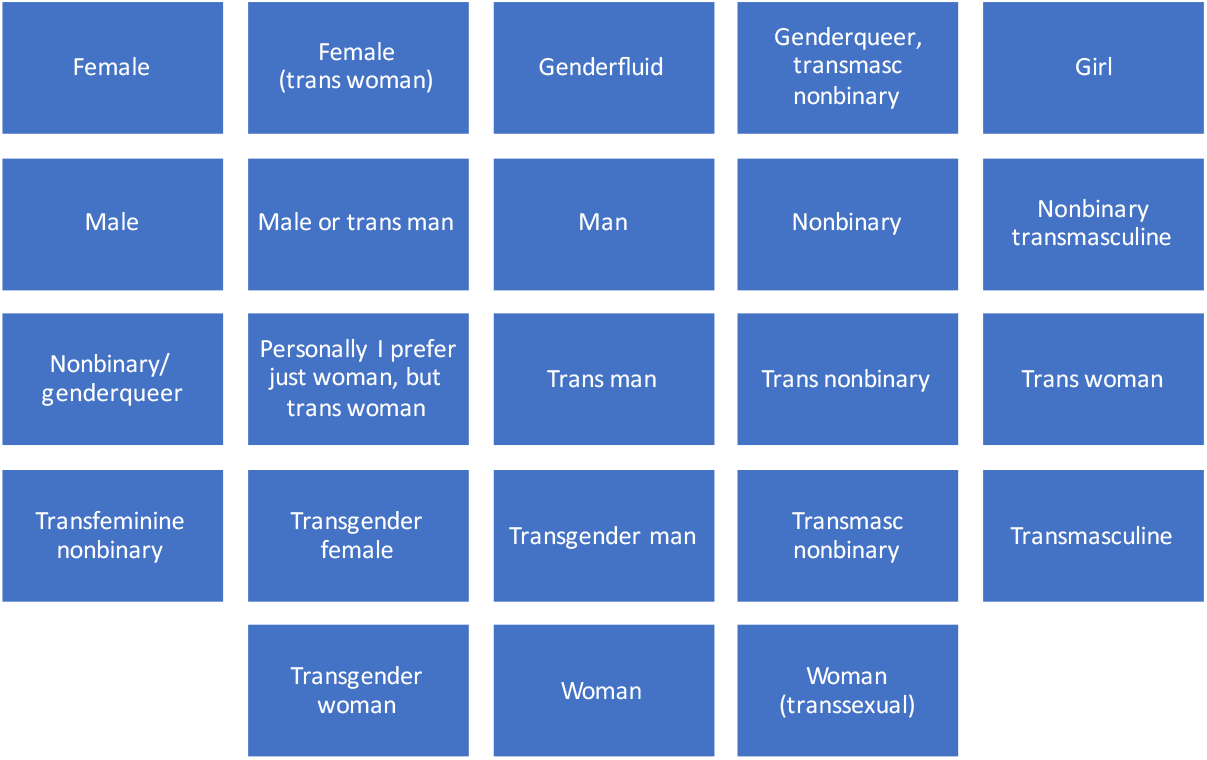
Participant self-described gender.

## Data Availability

All data produced in the present study are available upon reasonable request to the authors

## Notes

### Competing Interest Statement

The authors have declared no competing interest.

### Funding Statement

This study is funded by the National Institute of Health and Care Research, HPRU grant (NIHR200911)

### Author Declarations

Ethical approval was granted by University College London Research Ethics Committee (8805/007)

## References

1. Vincent BW. Studying trans: recommendations for ethical recruitment and collaboration with transgender participants in academic research. Psychology & Sexuality. 2018;9(2):102–16.

2. (ONS) OfNS. Gender identity, England and Wales: Census 2021 6 January 2023 [

3. McKechnie DGJ, O’Nions E, Bailey J, Hobbs L, Gillespie F, Petersen I. Transgender identity in young people and adults recorded in UK primary care electronic patient records: retrospective, dynamic, cohort study. BMJ medicine. 2023;2(1).

4. Stonewall. LGBT in Britain: The trans report.

5. Jones S, Patel T. Inaccessible and stigmatizing: LGBTQ+ youth perspectives of services and sexual violence. Journal of LGBT Youth. 2023;20(3):632–57.

6. Klein A, Golub SA. Family Rejection as a Predictor of Suicide Attempts and Substance Misuse Among Transgender and Gender Nonconforming Adults. LGBT Health. 2016;3(3):193–9.

7. Bockting WO, Miner MH, Romine RES, Hamilton A, Coleman E. Stigma, Mental Health, and Resilience in an Online Sample of the US Transgender Population. American Journal of Public Health. 2013;103(5):943–51.

8. Snow A, Cerel J, Loeffler DN, Flaherty C. Barriers to Mental Health Care for Transgender and Gender-Nonconforming Adults: A Systematic Literature Review. Health & Social Work. 2019;44(3):149–55.

9. Pezaro S, Crowther R, Pearce G, Jowett A, Godfrey-Isaacs L, Samuels I, et al. Perinatal Care for Trans and Nonbinary People Birthing in Heteronormative “Maternity” Services: Experiences and Educational Needs of Professionals. Gender & Society. 2023;37(1):124–51.

10. Connolly D, Hughes X, Berner A. Barriers and facilitators to cervical cancer screening among transgender men and non-binary people with a cervix: A systematic narrative review. Preventive Medicine. 2020;135:106071.

11. Mills TJ, Riddell KE, Price E, Smith DRR. ‘Stuck in the System’: An Interpretative Phenomenological Analysis of Transmasculine Experiences of Gender Transition in the UK. Qualitative Health Research. 2023;33(7):578–88.

12. Davy Z, Benson J, Barras A. Shared care and gender identity support in Primary Care: The perspectives and experiences of parents/carers of young trans people. Health. 0(0):13634593221138616.

13. Saadat M, Keramat A, Jahanfar S, Nazari AM, Ranjbar H, Motaghi Z. Barriers and Facilitators to Accessing Sexual and Reproductive Health Services Among Transgender People: A Meta-Synthesis. International Journal of Social Determinants of Health and Health Services. 0(0):27551938231187863.

14. Poteat T, Scheim A, Xavier J, Reisner S, Baral S. Global Epidemiology of HIV Infection and Related Syndemics Affecting Transgender People. J Acquir Immune Defic Syndr. 2016;72 Suppl 3(Suppl 3):S210–9.

15. Jaspal R, Nambiar KZ, Delpech V, Tariq S. HIV and trans and non-binary people in the UK. Sexually Transmitted Infections. 2018;94(5):318–9.

16. Hibbert MP, Wolton A, Weeks H, Ross M, Brett CE, Porcellato LA, et al. Psychosocial and sexual factors associated with recent sexual health clinic attendance and HIV testing among trans people in the UK. BMJ Sexual & Reproductive Health. 2020;46(2):116–25.

17. Day S, Smith J, Perera S, Jones S, Kinsella R. Beyond the binary: sexual health outcomes of transgender and non-binary service users of an online sexual health service. International journal of STD & AIDS. 2021;32(10):896–902.

18. Kirwan P, Hibbert M, Kall M, Nambiar K, Ross M, Croxford S, et al. HIV prevalence and HIV clinical outcomes of transgender and gender-diverse people in England. HIV medicine. 2021;22(2):131–9.

19. Ogbonmwan D, Hussey J, Mitchell L. Evaluating the clinical experience of sexual health trainees in the management of transgender, including non-binary, people within sexual health services. Sexually Transmitted Infections. 2020;96(5):320–1.

20. Malterud K, Siersma VD, Guassora AD. Sample Size in Qualitative Interview Studies: Guided by Information Power. Qual Health Res. 2016;26(13):1753–60.

21. Braun V, Clarke V. Toward good practice in thematic analysis: Avoiding common problems and be(com)ing a knowing researcher. International Journal of Transgender Health. 2023;24(1):1–6.

22. Wall CSJ, Patev AJ, Benotsch EG. Trans broken arm syndrome: A mixed-methods exploration of gender-related medical misattribution and invasive questioning. Social Science & Medicine. 2023;320:115748.

23. Norris M, Borneskog C. The Cisnormative Blindspot Explained: Healthcare Experiences of Trans Men and Non-Binary Persons and the accessibility to inclusive sexual & reproductive Healthcare, an integrative review. Sexual & Reproductive Healthcare. 2022:100733.

24. Dawson-Rose C, Cuca YP, Webel AR, Solís Báez SS, Holzemer WL, Rivero-Méndez M, et al. Building Trust and Relationships Between Patients and Providers: An Essential Complement to Health Literacy in HIV Care. Journal of the Association of Nurses in AIDS Care. 2016;27(5):574–84.

25. Dinç L, Gastmans C. Trust and trustworthiness in nursing: an argument-based literature review. Nursing Inquiry. 2012;19(3):223–37.

26. McPhail D, Lorway R, Chevrier C. Safe spaces that matter: Material semiotics, affective bodies and queer readings of clinical spaces in Winnipeg, Canada. SSM - Qualitative Research in Health. 2022;2.

27. Gilmore JP, Dainton M, Halpin N. Authentic allyship for gender minorities. Journal of Nursing Scholarship. 2024;56(1):5–8.

28. Brook G, Church H, Evans C, Jenkinson N, McClean H, Mohammed H, et al. 2019 UK national guideline for consultations requiring sexual history taking: clinical effectiveness group British Association for Sexual Health and HIV. International journal of STD & AIDS. 2020;31(10):920–38.

29. Gruskin S, Everhart A, Olivia DF, Baral S, Reisner SL, Kismödi E, et al. “In transition: ensuring the sexual and reproductive health and rights of transgender populations.” A roundtable discussion. Reproductive Health Matters. 2018;26(52):21–32.

30. Mikulak M, Ryan S, Ma R, Martin S, Stewart J, Davidson S, et al. Health professionals’ identified barriers to trans health care: a qualitative interview study. British Journal of General Practice. 2021;71(713):e941–e7.

